# Sars-Cov-2 antibody titer 3 months post-vaccination is affected by age, gender, smoking and vitamin D

**DOI:** 10.1101/2021.09.01.21262913

**Authors:** Anastasia Parthymou, Evagelia E Habeos, George I Habeos, Apostolos Deligakis, Ektoras Livieratos, Markos Marangos, Dionysios V Chartoumpekis

## Abstract

**Context:** Vaccination against Sars-Cov-2 is in full swing during COVID-19 pandemic. One of the efficient methods to evaluate response to vaccination is the assessment of humoral immunity by measuring Sars-Cov-2 antibody titer. Identification of factors that affect the humoral response is important so as to ameliorate the responses to vaccination or identify vulnerable groups that may need vaccination boosters.

**Objective:** We investigated the effect of anthropometric parameters (age, BMI), smoking, diabetes, statin use hypertension and levels of 25(OH)D and DHEAS to the Sars-Cov-2 antibody titer.

**Methods:** In this longitudinal observational cohort study 712 subjects were tested for Sars-Cov-2 antibodies 3 months after the second dose of BNT162b2 vaccine. Multiple linear regression analysis was performed to identify which factors are associated with the antibody titer.

**Results:** We identified age to be negatively associated with antibody titer (p=0.0073) and male sex (p=0.0008). However, interaction of age and gender was significant (p<0.0001) highlighting the finding that only after the age of 40 years men had lower antibody levels than women. DHEAS, an aging marker, was not associated with the antibody titer. Smoking was also associated with low antibody titer (p=0.0008) while overweight or obese subjects did not have different antibody response compared to normal weight individuals. Although diabetic and hypertensive subjects trended towards lower antibody titer, this association was not statistically significant. Replete vitamin D levels were associated with higher antibody titers (p=0.00422).

**Conclusions:** Age, male sex and smoking negatively affects antibody titer while 25(OH)D is associated with increased Sars-Cov-2 antibody titers.

## Introduction

SARS-CoV-2 has spread worldwide since January 2020 resulting by the end of July 2021 to about 197 million confirmed cases of COVID-19 with over 4 million deaths according to World Health Organization. Pharmaceutical and biotechnology companies started developing COVID-19 vaccines and four have received so far authorization to be used in European Union (BioNTech and Pfizer, Moderna, AstraZeneca, Johnson & Johnson/Janssen Pharmaceuticals). BNT162b2 vaccine developed by BioNTech and Pfizer is a lipid nanoparticle-formulated nucleoside-modified RNA encoding the SARS-CoV-2 spike glycoprotein (1) and was the first to receive approval for use in European Union, hence being of the first to be administered in Greece. The safety and 95% efficacy in preventing symptomatic COVID-19 of BNT162b2 vaccine had been initially shown by the BioNtech-Pfizer clinical trials(1) and similar results were obtained in other studies(2-5).

The humoral immune response to COVID-19 vaccination has usually been assessed in clinical trials by measuring the antibody titer (including IgG) against the SARS-CoV-2 spike (S) protein(6,7). However, FDA and other regulatory bodies do not recommend assessment of immunity after COVID-19 vaccination by measuring antibody titer. Nevertheless, commercial serological assays show over 90% positive agreement for distinguishing neutralizing antibodies at the same neutralizing titer while the negative percent agreement was poorer (less than 80%) (8). Hence, antibody titer against SARS-CoV-2 spike (S) protein can be an easy measure of the humoral immune response to COVID-19 vaccination in the real world.

It has been a matter of concern that specific populations will not be able to develop an effective immune response after COVID-19 vaccination. Such groups include but they are not limited to aged individuals, immunocompromised patients due to autoimmune diseases or due to immunosuppressive medication, cancer patients and obese/diabetic patients. Thus, studies with kidney transplant recipients (9), cancer patients(10,11) came into focus. Moreover, the identification of other factors that can possibly impair or boost the immune response is important so as to identify individuals that might be at risk even after COVID-19 vaccination. Such factors can be obesity (12), aging (13), dehydroepiandrosterone sulfate (DHEAS), a steroid that declines with aging (14), vitamin D (15), statin use (16), smoking (17) and hypertension (18).

As previous studies on SARS-CoV-2 antibodies response (reviewed in (19)) are limited by number of patients and their methodologies, we report herein the real-world humoral immunogenic response to BNT162b2 vaccine in subjects of western Greece vaccinated against COVID-19, and we assess potential parameters that may affect this response.

## Methods

### Design, setting and study population

This is a longitudinal observational cohort study (ClinicalTrials.gov Identifier: NCT04954651, approved by the University Hospital of Patras Ethics Committee, approval ID 99-25/2/202) in healthcare units of western Greece. Blood samples were drawn before vaccination, 3 weeks and 3 months after the second dose of vaccination with BNT162b2 vaccine (BioNTech and Pfizer) as part of the national COVID-19 vaccination program in Greece. Blood samples at the 3-weeks timepoint were drawn only from a portion of our cohort (**Fig. 1**). Participation was based on informed written consent. All adults of any age and gender were eligible to participate in this study. Data about body weight, height, smoking, diabetes, hypertension and statin use were obtained from all participants based on questionnaires. Exclusion criteria were: past medical history of COVID-19 or infection with SARS-Cov-2 during the study or positive SARS-CoV-2 spike (S) protein antibody titer at baseline (before vaccination). Moreover, patients who did not provide all necessary information in the questionnaire or there was not enough sample to run all the assays were excluded from our analyses (**Fig. 1**). As healthcare personnel was prioritized for vaccination in Greece, our cohort consists to a large degree of subjects working at hospitals or health centers.

**Figure 1.**
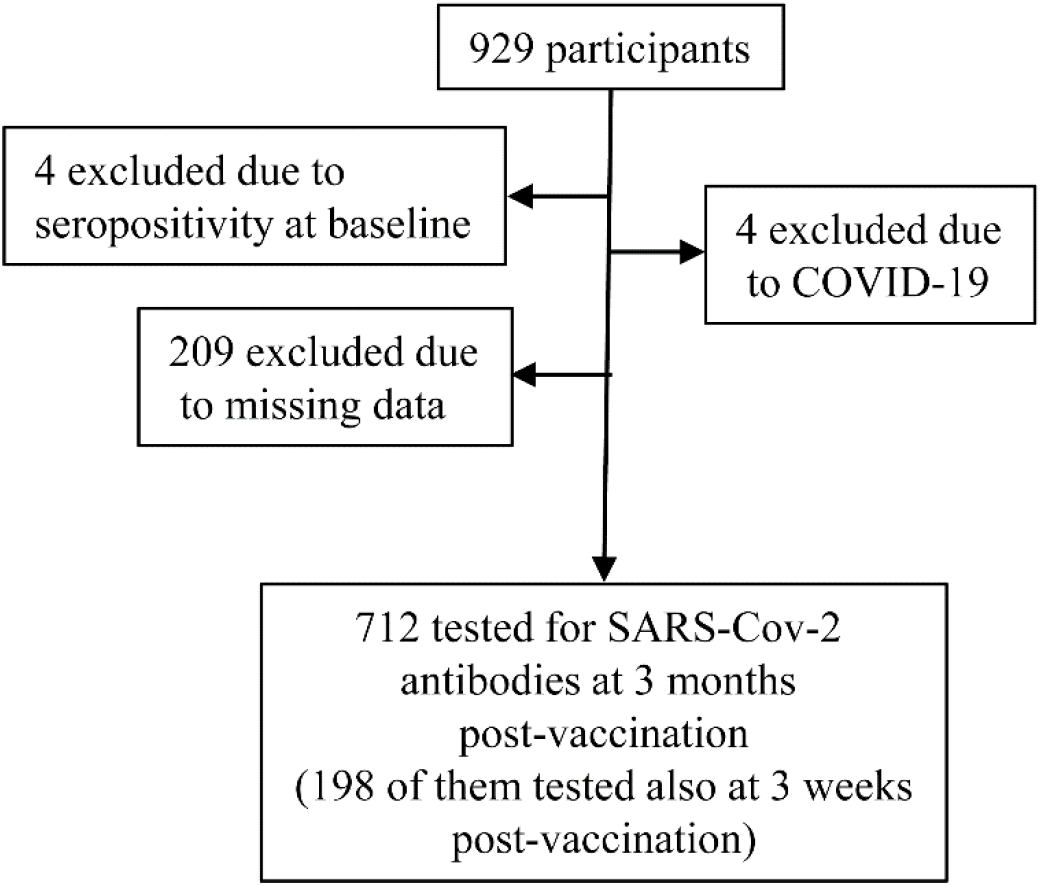
Study profile. “Post-vaccination” means after the second dose of the BNT162b2 vaccine. Missing data means that patients did not provide all necessary information in the questionnaire or there was not enough sample to run all the assays.

**Figure 2.**
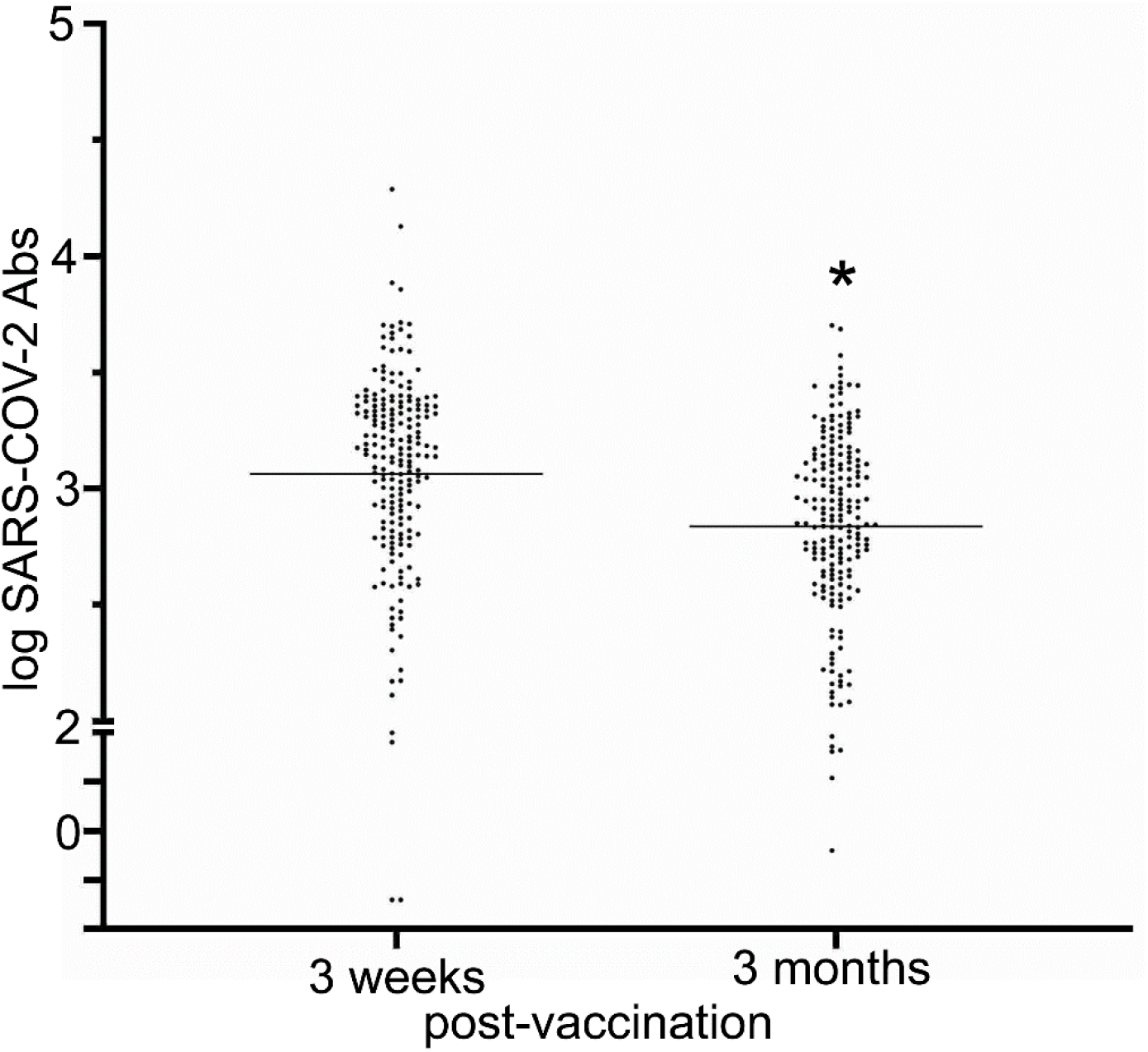
SARS-COV-2 antibody titer from 3 weeks to 3 months post-vaccination with BNT162b2.Dots indicate individual values (n=198) and straight line indicates means of log antibody titer. *p<0.0001, paired t-test.

### Laboratory measurements

Serum samples were stored in a -20°C freezer. All serum samples measurements were performed in a Cobas 6000 E601 analyzer (Roche Diagnostics GmbH, Mannheim, Germany) using electrochemiluminescence (ECL) technology for immunoassay analysis. Quantitative determination of antibodies (including IgG) to SARS CoV 2 spike (S) protein receptor binding domain (RBD) was performed using the Elecsys^®^ Anti-SARS-CoV-2 S immunoassay (Roche). Antibody levels higher than 0.8 U/l were considered positive (measurement range 0.4-250 U/ml). Values higher than 250 U/ml were diluted 1:20 to obtain the exact value. 25(OH)D (vitamin D) and DHEAS levels were assessed at the 3-months post-vaccination timepoint. The measuring range for DHEAS is 0.2-1000 μg/dl and for Vitamin D is 3.00-120 ng/mL respectively.

### Statistical analysis

Descriptive statistics for baseline characteristics of the study population were calculated as means ± standard deviation (SD). To assess which factors affect the SARS-Cov-2 antibody titer, multiple linear regression modelling was performed with log-transformed antibody titer as a continuous dependent variable and using stepwise entry criteria of p<0.05. Age, sex, smoking, hypertension, diabetes, statin use, body mass index (BMI), DHEAS and Vitamin D were checked for linear association with the log-transformed antibody titer and if no linear association was found for a variable, this variable was converted to categorical using quartiles (DHEAS and vitamin D) or pre-existing categories for BMI (underweight; <18.5, normal weight; 18.5-24.9, overweight; 25-29.9, obesity; >30). Participants who did not answer all of the questions from the questionnaire were excluded from the analysis. GraphPad Prism version 9.0.0 for Windows (GraphPad Software, San Diego, CA, USA) was used for all the statistical analyses and graphs generation. **Fig.3** and **Fig.4** graphs were generated using Microsoft Excel 365 (Microsoft, Redmond, WA, USA).

**Figure 3.**
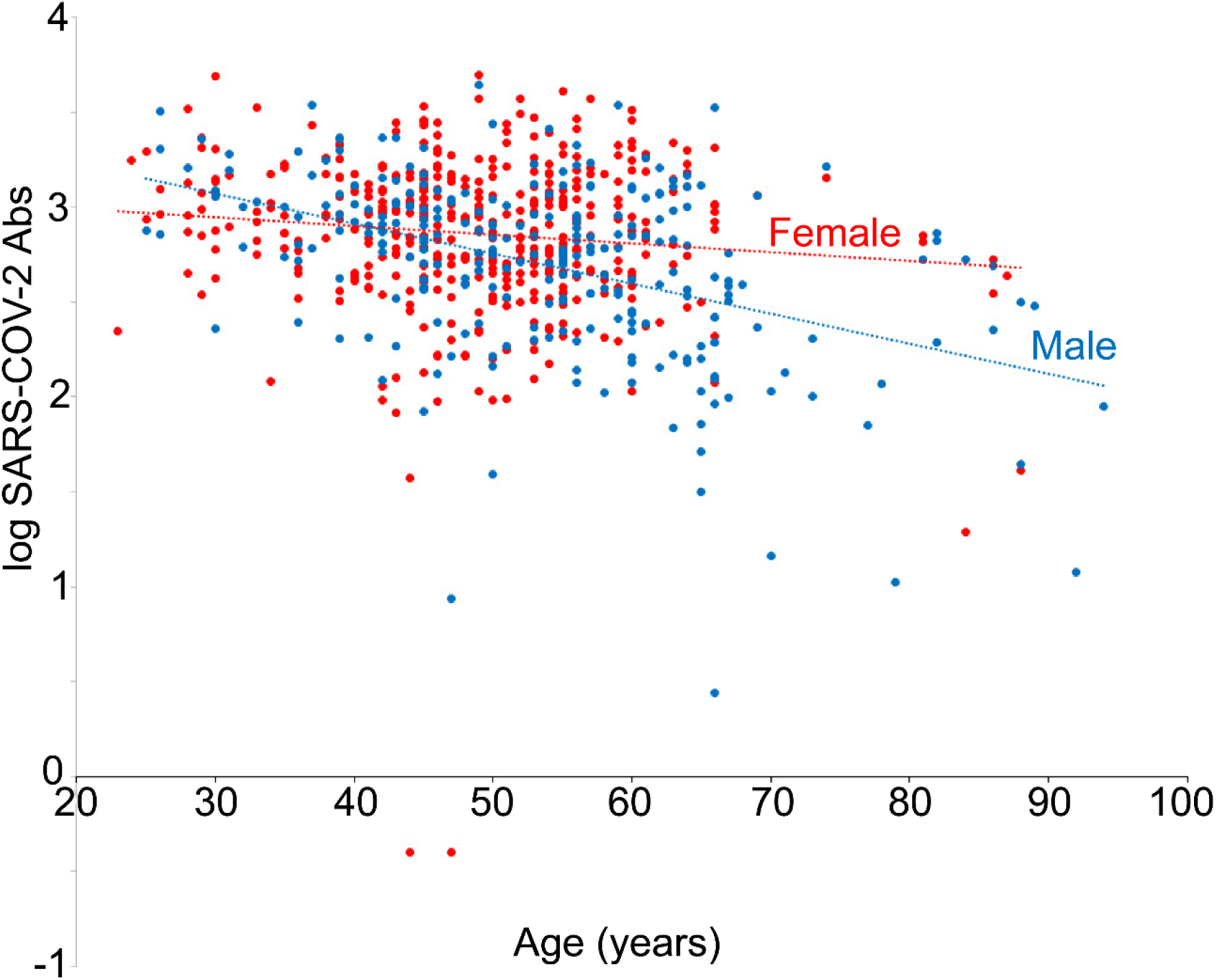
SARS-COV-2 antibody titer at 3 months post-vaccination plotted against age (n=712). Spearman’s r=-0.247, p<0.0001. Red dots indicate female gender and blue dots male gender.

**Figure 4.**
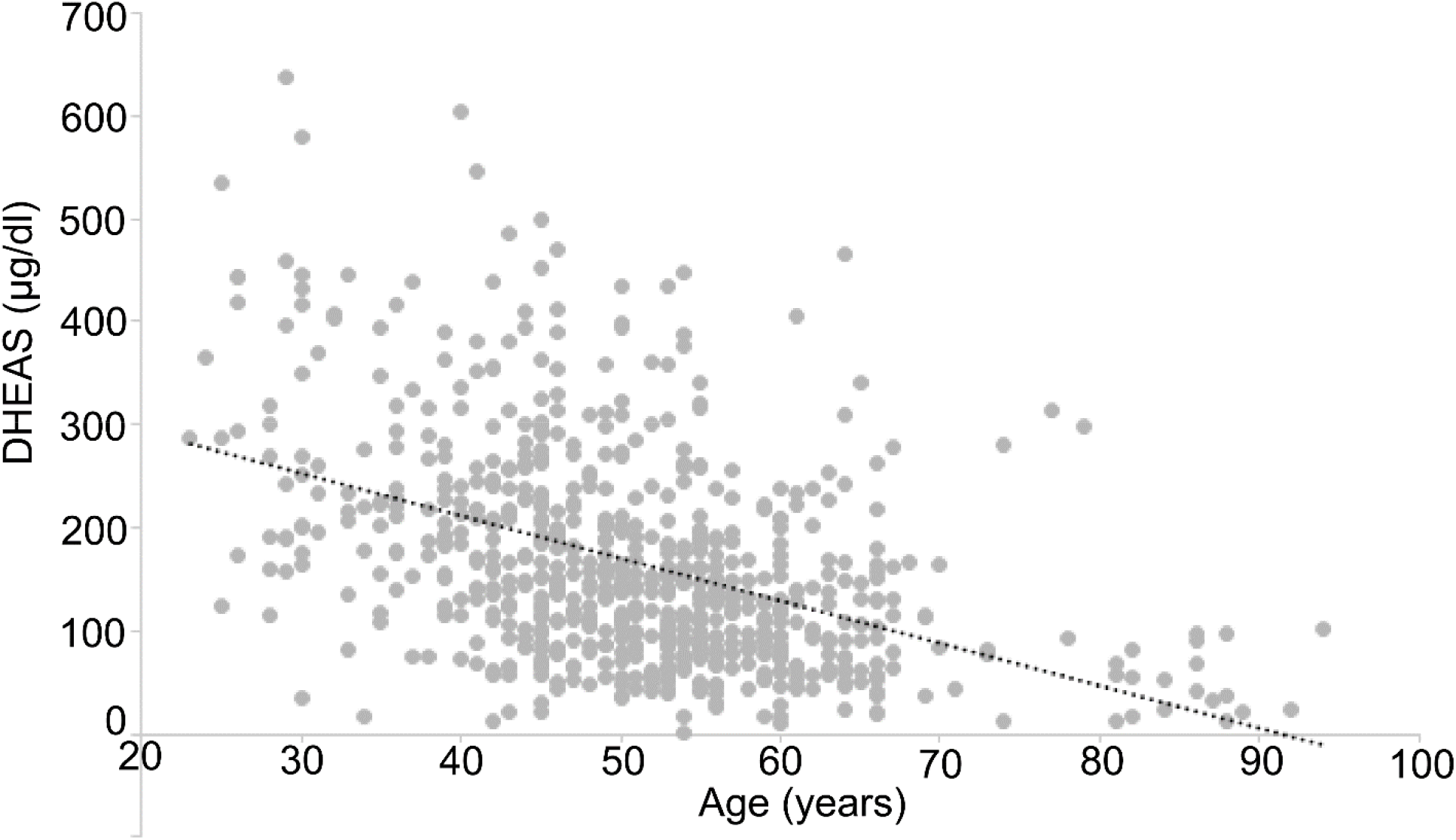
DHEAS levels plotted against age at 3 months post-vaccination. Spearman’s r=-0.46, p<0.0001

## Results

### Characteristics of the study sample

Of the 929 participants in the present study, 8 were excluded due to being tested positive for SARS-COV-2 antibodies at baseline or suffered from COVID-19 during the study (**Fig.1**). 209 participants were also excluded because at least one parameter in the questionnaire was not answered (for instance not giving information about their age, their body weight etc) or due to technical reasons (not enough sample to run the assay). The serum of the remaining 712 participants was assayed for SARS-COV-2 antibodies, DHEAS and vitamin D, 3 months post-vaccination. 198 of them were also tested for SARS-COV-2 antibodies 3 weeks after the second dose of the vaccine (**Fig. 1**). The baseline characteristics (gender, age, BMI, smoking, diabetes, hypertension, use of statins) of our study population (n=712) can be seen in **Table 1**.

**Table 1.** Baseline characteristics of the study population.

| Whole study population | n=712 |
| --- | --- |
| Male | 268 (37.6%) |
| Female | 444 (62.4%) |
| Age (years) | 50.8 ± 11.4 |
| BMI (kg/m<sup>2</sup>) | 26.7 ± 4.9 |
| Smokers | 245 (34.4%) |
| Diabetes | 50 (7%) |
| Hypertension | 115 (16.2%) |
| Statin use | 108 (15.2 %) |

Data show means ± SD

### SARS-COV-2 antibodies decline from 3 weeks to 3 months post-vaccination

Paired serum samples from a part of cohort (n=198) were tested at 3 weeks as well as 3 months post-vaccination with the second dose of BNT162b2 vaccine. The SARS-COV-2 antibodies levels were assessed at both of these timepoints. Pairing was significantly effective (correlation coefficient r=0.8139, p<0.0001) and paired t-test showed that the log antibody titer significantly declines with time (mean of differences between 3 months and 3 weeks is -0.23 ± 0.34 (p<0.0001) (**Fig. 2**).

### Multiple linear regression analysis of the effect of various factors on SARS-COV-2 antibody titer

To examine which factors can potentially affect the SARS-COV-2 antibody titer in our cohort 3 months post-vaccination, we employed a multiple linear regression analysis model, including initially all considered parameters (gender, age, smoking, hypertension, hypertension, diabetes, statin use, BMI, vitamin D and DHEAS levels). As BMI, DHEAS and vitamin D levels did not show linear association with the antibody titer, these variables were converted to categorical ones using quartiles for DHEAS and Vitamin D, and the categories of underweight, normal weight, overweight and obese in the case of BMI. The results of the multiple linear regression analysis model that incorporates all these factors can be seen in **Table 2**. Using step-wise entry criteria with p<0.05, the parameters diabetes, hypertension, prior statin use, BMI and DHEAs were excluded one by one with the aforementioned order rebuilding each time the linear regression model. Finally, the parameters that were found to affect significantly the antibody titer were age, gender, smoking and vitamin D levels (**Table 3**).

**Table 2.** Initial multivariable linear regression model for SARS-COV-2 antibodies titer

| Variable | Beta (95% CI) | p-value |
| --- | --- | --- |
| Gender (male) | -0.1041 (-0.1747 to -0.0335) | 0.0039 |
| Age | -0.01300 (-0.0164 to -0.0096) | <0.0001 |
| Smoking (yes) | -0.1006 (-0.1653 to -0.03599) | 0.0023 |
| Hypertension (yes) | -0.04540 (-0.1364 to 0.04560) | 0.3276 |
| Diabetes (yes) | -0.01871(-0.1433 to 0.1059) | 0.7681 |
| Statin (yes) | 0.06516 (-0.02489 to 0.1552) | 0.1559 |
| BMI (underweight) | -0.1507 (-0.4253 to 0.1239) | 0.2816 |
| BMI (overweight) | 0.08446 (0.01126 to 0.1577) | 0.0238 |
| BMI (obese) | 0.06558 (-0.01823 to 0.1494) | 0.1249 |
| DHEAS (Q3) | -0.03468 (-0.1218 to 0.05248) | 0.4350 |
| DHEAS (Q2) | -0.03305 (-0.1244 to 0.05827) | 0.4776 |
| DHEAS (Q1) | -0.1229 (-0.2226 to -0.02312) | 0.0158 |
| 25(OH)D (Q3) | 0.08445 (-0.001423 to 0.1703) | 0.0539 |
| 25(OH)D (Q2) | 0.1396 (0.05289 to 0.2262) | 0.0016 |
| 25(OH)D (Q1) | 0.1158 (0.02801 to 0.2035) | 0.0098 |

25(OH)D quartiles: Q4; 4.1-18.89, Q3; 18.9-25.67, Q2; 25.68-32.99 Q1; 33-69.8 ng/ml
DHEAS quartiles: Q4; 0.3-89.59, Q3; 89.6-145.9, Q2; 146-217.39, Q1; 217.4-637.6 µg/dl

**Table 3.** Final multivariable linear regression model for SARS-COV-2 antibodies titer

| Variable | Beta (95% CI) | p-value |
| --- | --- | --- |
| Gender (male) | -0.1100 (-0.1738 to -0.04617) | 0.0008 |
| Age | -0.005231 (-0.009047 to -0.001416) | 0.0073 |
| Smoking (yes) | -0.1097 (-0.1737 to -0.04567) | 0.0008 |
| 25(OH)D (Q3) | 0.07601 (-0.009362 to 0.1614) | 0.0809 |
| 25(OH)D (Q2) | 0.1231(0.03724 to 0.2091) | 0.0050 |
| 25(OH)D (Q1) | 0.08248 (-0.003949 to 0.1689) | 0.0614 |
| Gender (male):age | -0.01090 (-0.01631 to -0.005490) | <0.0001 |

Adjusted R<sup>2</sup>=0.1355
25(OH)D quartiles: Q4; 4.1-18.89, Q3; 18.9-25.67, Q2; 25.68-32.99 Q1; 33-69.8 ng/ml
25(OH)D as categorical data Q1, Q2, Q3 compared to Q4 show p=0.0422

### Influence of age and gender on SARS-COV-2 antibody titer 3 months post-vaccination

Multivariate linear regression analysis highlighted a statistically significant decreased response of the antibody titer with increasing age (**Table 3, Fig.3**). Male gender also appears to have a negative effect on the antibody titer (**Table 3**). As it is shown in **Fig. 3** male subjects tend to have lower antibody titer compared to female after the age of 40. Indeed, the effect of gender on antibody titer is modified by age as the interaction of sex and age is significant (p<0.0001, **Table 3**).

Since DHEAS has been reported as an aging marker with its concentration negatively correlated with age, we hypothesized that DHEAS levels might predict SARS-COV-2 antibody titers. Indeed, in our cohort we confirmed the known negative association between age and DHEAS level (Spearman’s r=-0.46, p<0.0001, **Fig. 4**) but multivariate linear regression analysis did not find DHEAS to be a significant predictor of antibody titer.

### Metabolic parameters and SARS-COV-2 antibody response 3 months post-vaccination

The metabolic profile of our cohort included information from questionnaires about BMI, history of hypertension or use of antihypertensive drugs, history of diabetes and use of statins. Overweight and obese individuals did not show any different antibody response to vaccination compared to normal weight subjects (**Fig. 5, Table 2)**. Even though individuals with diabetes or hypertension tended to have lower levels of antibodies (**Fig. 6** and **Fig.7** respectively) multivariate linear regression analysis showed that these variables do not affect antibody titer independently (**Table 2**). Users of statin treatment were not found to have different antibody response compared to non-users (**Table 2**).

**Figure 5.**
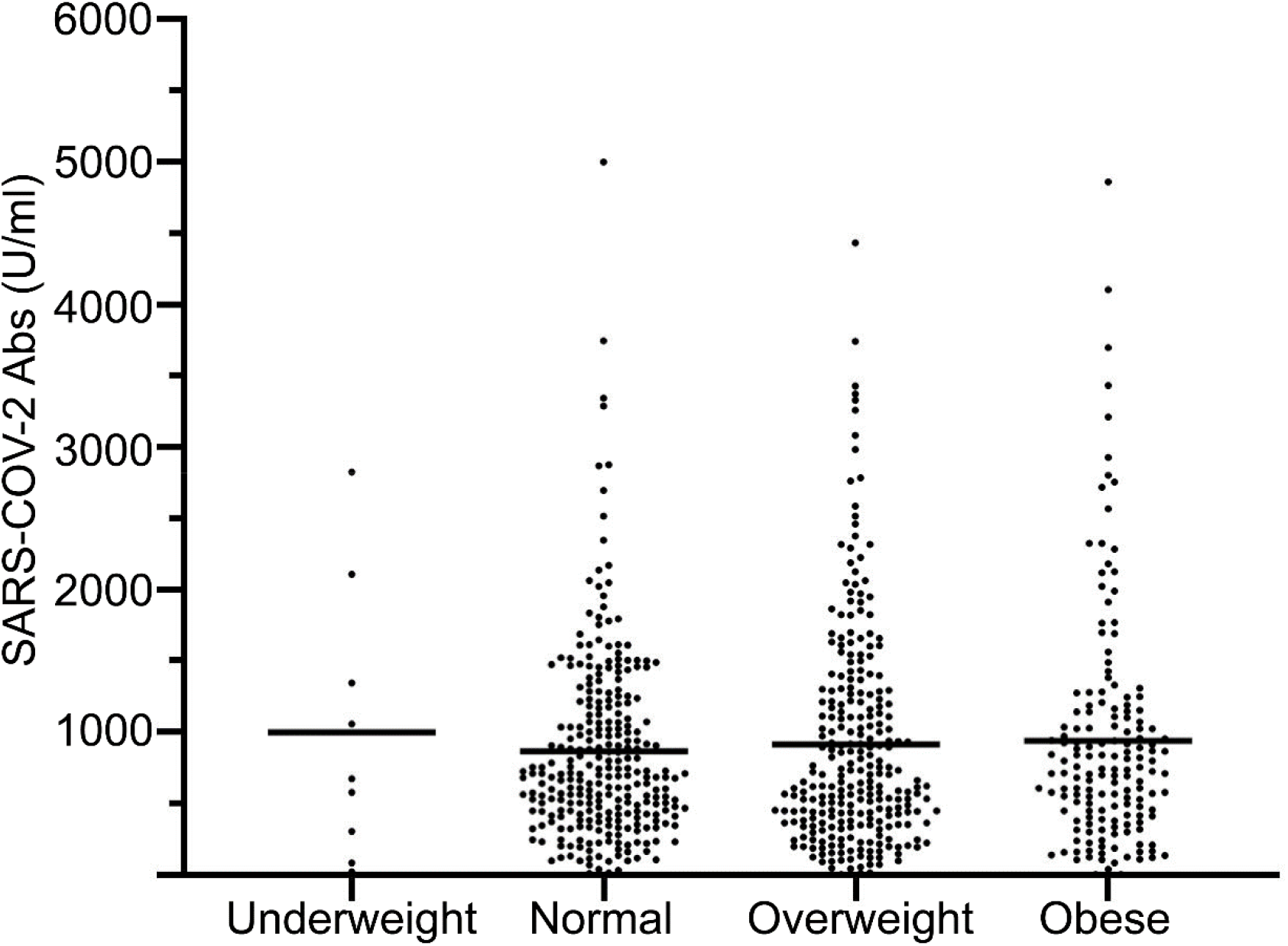
SARS-COV-2 antibody titer in underweight (BMI<18.5), normal weight (BM=18.5-24.9), overweight (BMI=25-29.9) and obese (BMI>30) subjects at 3 months post-vaccination. Individual values are depicted with the straight line indicating the means.

**Figure 6.**
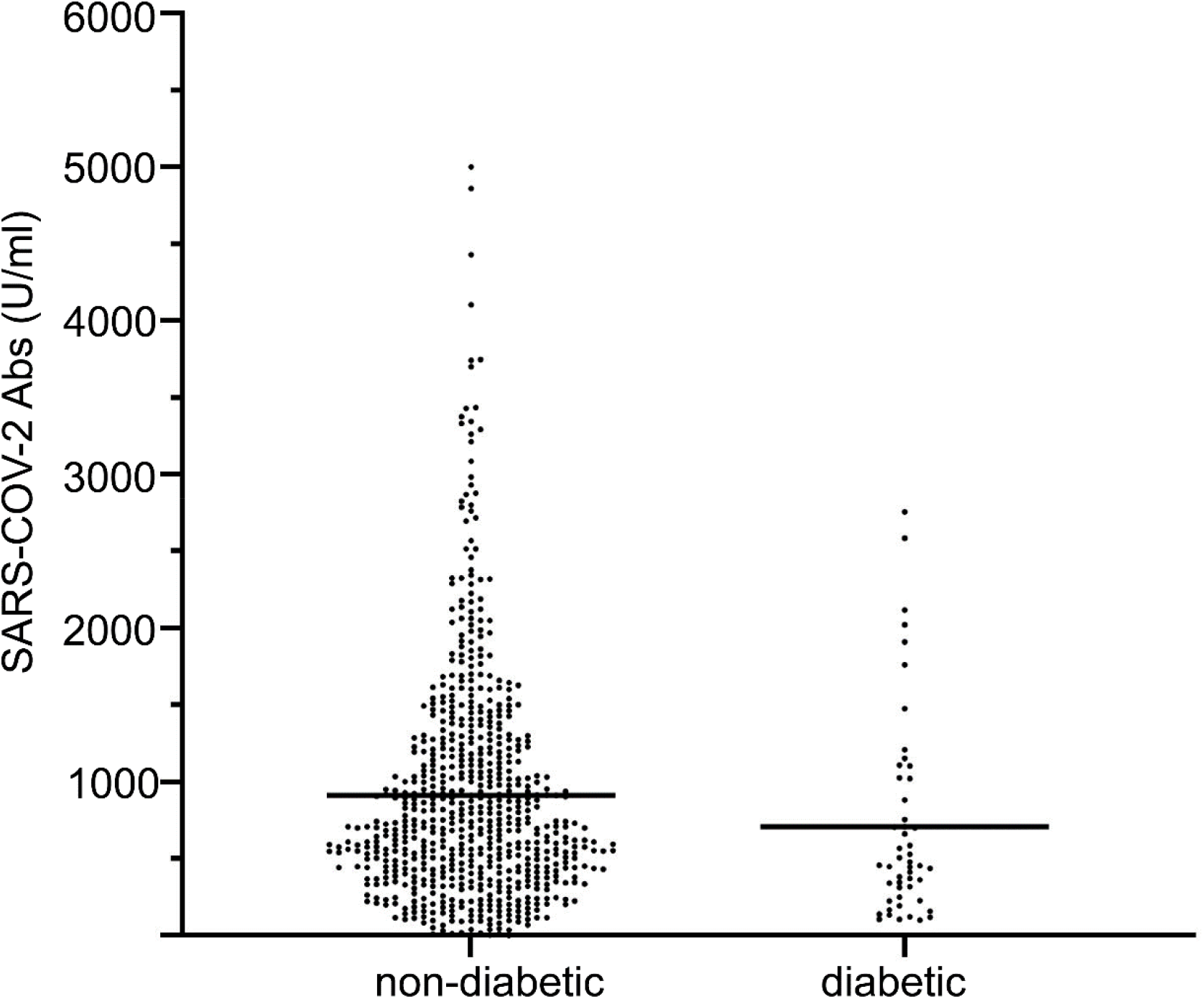
SARS-COV-2 antibody titer in diabetic and non-diabetic subjects at 3 months post-vaccination. The presence of diabetes was based on the reported treatment of diabetes medication by the subjects in the questionnaires. Individual values are depicted with the straight line indicating the means.

**Figure 7.**
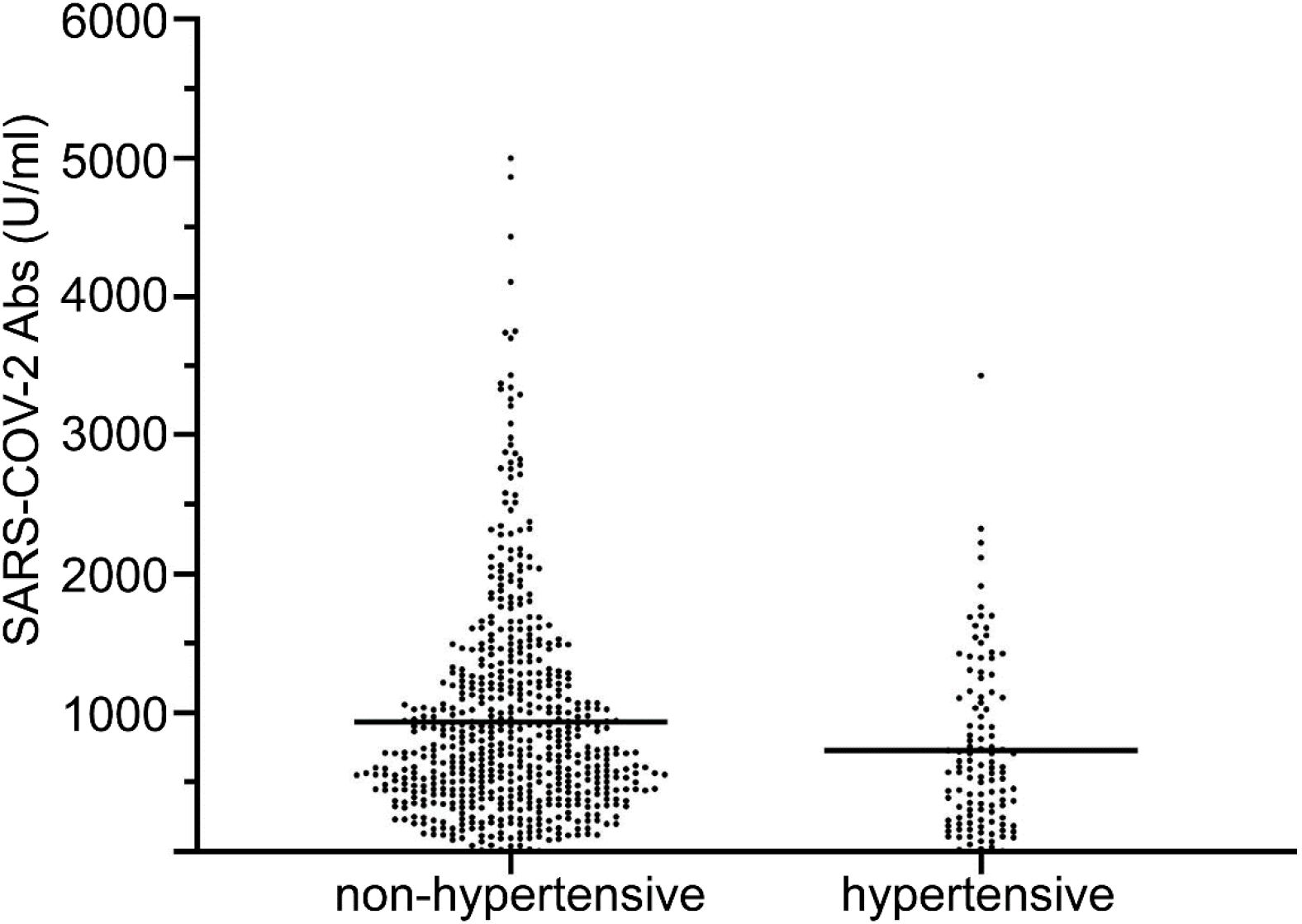
SARS-COV-2 antibody titer in hypertensive and non-hypertensive subjects at 3 months post-vaccination. The presence of hypertension was based on the reported treatment of antihypertensive medication by the subjects in the questionnaires. Individual values are depicted with the straight line indicating the means.

### Smokers showed a blunted humoral response to vaccination

In our cohort 245 out of 712 subjects (34.4%) were smokers. Multivariate linear regression analysis revealed a negative association between smoking and antibody titer (95% CI -0.1737 to -0.04567, p=0.0008, **Table 3**). The mean antibody titer of smokers 988±781.4 versus 731.2±603.9 in non-smokers (**Fig. 8**).

**Figure 8.**
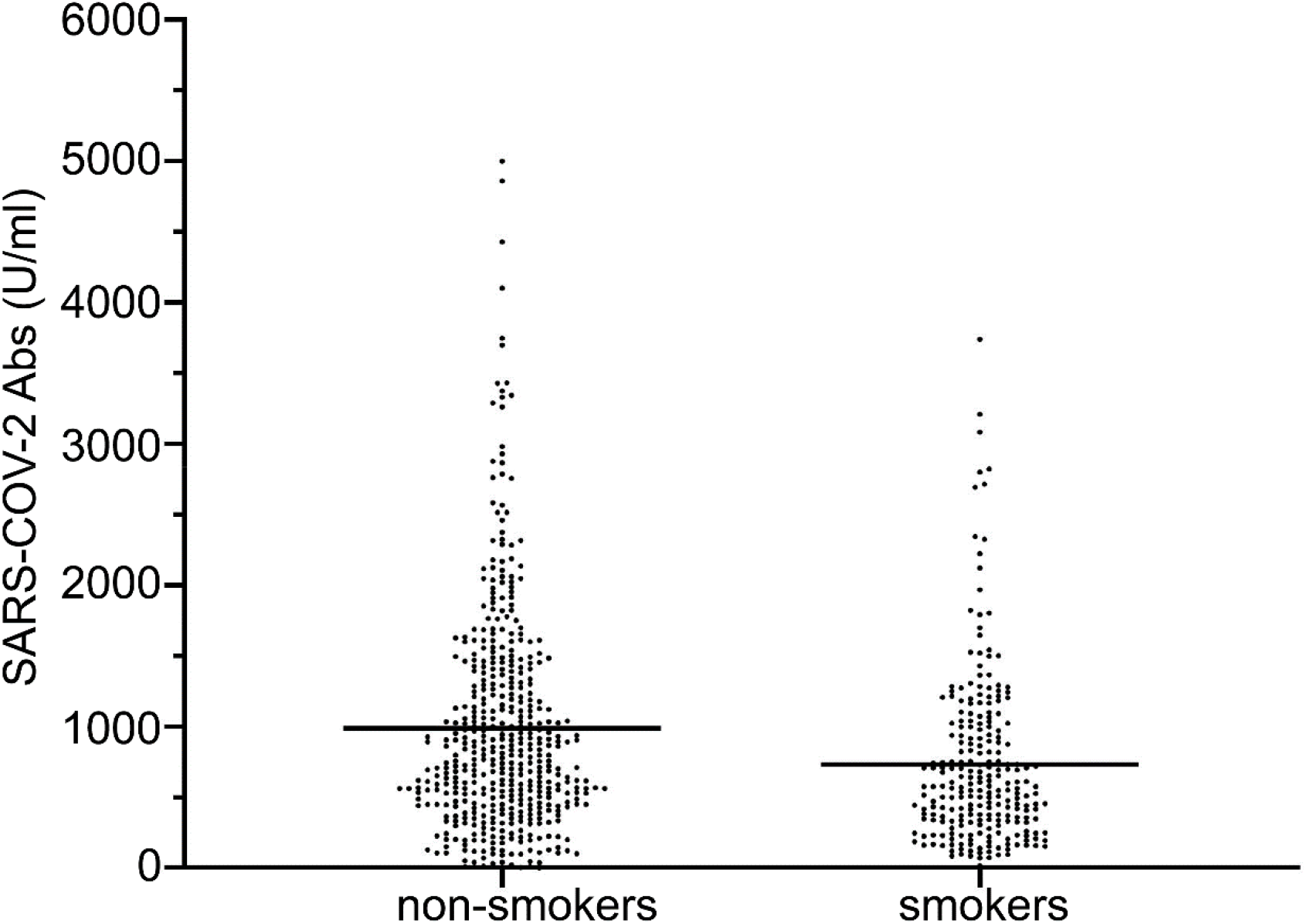
SARS-COV-2 antibody titer in smokers and non-smokers subjects at 3 months post-vaccination. Individual values are depicted with the straight line indicating the means. Multiple linear regression analysis showed statistically significant negative association (p=0.0008)

### Vitamin D status and humoral response to vaccination

The association of vitamin D and antibody titer was not linear and thus quartiles (Q4; 4.1-18.89, Q3; 18.9-25.67, Q2; 25.68-32.99 Q1; 33-69.8 ng/ml) of vitamin D levels were used instead in the multivariate linear regression model. Q1, Q2 and Q3 quartiles were compared to Q4. A statistically significant association was found between vitamin D levels and antibody response (p=0.0422, **Table 3**). Specifically, individuals with vitamin D levels in the Q2 quartile showed a positive association with antibody titer (95% CI 0.03724 to 0.2091, p=0.0050) while Q1 and Q3 quartiles trended to have positive association (**Fig.9, Table 3**).

**Figure 9.**
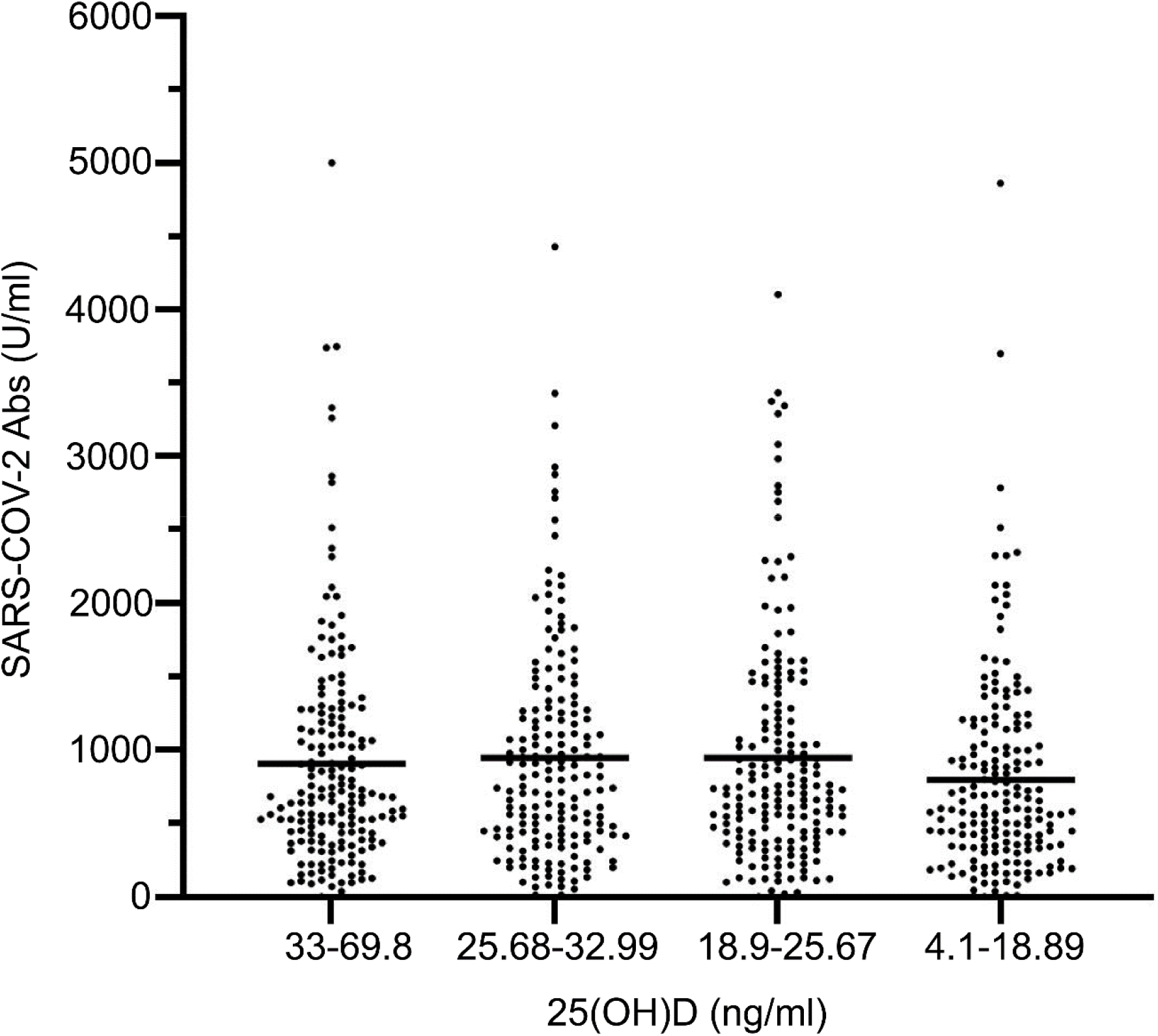
SARS-COV-2 antibody titer in subjects with different levels of vitamin D. Individual values are depicted with the straight line indicating the means. Multiple linear regression analysis showed statistically significant positive association with statistically significant effect on the vitamin D levels range 25.68-32.99 ng/ml compared to 4.1-18.89 ng/ml.

## Discussion

In the present work the potential association of SARS-COV-2 antibody titers after vaccination in volunteers consisting mainly of healthcare workers, was tested with a variety of potential parameters. Age was found to be negatively associated with the antibody titers and men over 40 years old maintained lower antibodies titers than women of the same age (**Fig. 3**). Aging has long been considered a factor that affects immunity (20) and has been recognized as an important factor in the current coronavirus pandemic (21). Aged individuals have shown higher mortality rates from COVID-19 (22). The lower antibody titer of aged subjects after vaccination may suggest an impaired immune response and it can be a matter of consideration for providing the aged population with booster shots. Another interesting finding is that males appear to have lower antibody levels but this effect is age-dependent and it is more obvious after the age of 40 (**Fig. 3**). This finding is in accordance with another study (23) that used a smaller number of subjects but assessed antibody levels in various timepoints from day 1 to day 50 post-vaccination. It is shown that octogenarians generate much less antibodies and that females older than 80 years old show higher titers. Other major studies detected a correlation of lower antibody concentrations after vaccination with male sex and older age (24,25). It is also noteworthy that age is negatively associated with antibody titer after Sars-Cov-2 infection (26). As men also appear to dominate in COVID-19 case fatality at least in some countries (reviewed in (27)), such findings warrant further investigation with focus on the mechanisms male gender and aging can have such effects on immune response and COVID-19 outcomes.

In our cohort the prevalence of smokers was relatively high, i.e. 245/712 (34.4%). Smoking was also concluded to be associated with lower antibody titer after vaccination (**Fig.8**, **Table 2**). This finding is in agreement with a recent smaller Italian study (28) and with a larger Japanese study (25) examining the antibody response after COVID-19 vaccination. A meta-analysis of 19 clinical studies revealed that smokers have higher odds of COVID-19 disease progression compared to non-smokers (29) indicating that smoking is associated with probably impaired immune response to Sars-Cov-2 or that comorbidities associated with smoking can lead to such a result. The effects of cigarette smoking to immunity has been studied through the years and is usually associated with immunosuppression (30). Hence, health policies aiming to reduce the smoking population should be more thoroughly implemented not only for the cancer prevention and cardiovascular benefits but also for the amelioration in immune responses.

Obesity has been officially recognized as a disease in some countries (for instance, by the American Medical Association) as it is a recognized risk factor of cardiovascular disease (31), cancer (32), diabetes, hypertension, dyslipidemia and other comorbidities (33). During the current coronavirus pandemic, obesity has been recognized as a risk factor for greater COVID-19 severity (34). In our study the simple index of BMI was calculated based on information provided by the participants. Analysis showed that BMI was not associated with the total antibody titer (**Fig.5, Table 2**). This is in accordance with other studies (24,28). However, another index of central obesity, waist circumference, has been found to be inversely associated with antibody titer (28). Our study is limited by the fact that body weight and height were not measured but they were based on self-reporting of the participants. Diabetes (**Fig.6, Table 2**) and hypertension (**Fig. 7, Table 2**) did not also appear to have an effect on the antibody titer in the present study. Again, the presence of diabetes was not based on HbA1C or on a glucose measurement but only on the reporting of antidiabetic medications. No index of poorly or well-controlled diabetes was used, just the presence or absence of diabetes was recorded based on the medications received. Similarly, hypertension was recorded in the same manner. Metanalyses have shown that diabetes increases the risk for severe COVID-19 (35,36) while reports for hypertension have been more controversial (37) as they have not corrected for potential cofounders such as cardiovascular disease. A recent study (24) showed a negative association between antibody titers and hypertension or diabetes. However, this is not the case in our study where we show no association. This discrepancy could be attributed to differences in the size, age and self-reporting of the populations. Further studies are warranted with more objective evaluation not only of the presence or absence of diabetes or hypertension but also with evaluation of indexes of poor or well management (e.g., HbA1C, measurement of blood pressure on-site etc.).

In our cohort among the variables tested was the prior use of statins. Statin use was considered as a potential determinant of antibody response based on previous knowledge of pleiotropic effect of statins on humoral immunity after vaccination (16). Moreover, use of statins in hospitalized COVID-19 patients has been reported to be associated with reduced mortality (38). A metanalysis also supported this notion (39) but concluded that more and especially prospective studies are needed. With reference to the effect of statins on the humoral response, a report on influenza vaccination focusing mainly in old population showed that chronic statin use had an immunosuppressive effect (16). In our study, statins had no significant effect on the antibody levels post-vaccination (**Table 2**) but it should be noted that only 15.2 % of our cohort used statins (**Table 1**).

Besides the anthropometric measurements and past medical history based on the questionnaires, 25(OH)D and DHEAS levels were assessed in the subjects of our study. These hormones were selected for different reasons. DHEAS is an aging marker (40) and has been indirectly implicated in immune responses (41). Although we confirmed in our series the negative association of DHEAS levels with age (**Fig. 4**), we could not find any association of DHEAS with the antibody levels (**Table 2**). This means that the negative effect of age on antibody titers is not dependent on DHEAS levels and other mechanisms should be investigated.

Vitamin D has been in the spotlight of research for its pleiotropic actions besides bone effects for at least the last 2 decades (15). Its effects on immune response (42) attracted our attention as they may affect the antibody titers after COVID-19 vaccination. Specifically, vitamin D supplementation has been shown to increase antibodies production after influenza vaccination in an elderly population (43) whereas vitamin D deficiency did not have any association with the immunogenic response to influenza vaccination according to a metanalysis (44). Another study did not find any effect of vitamin D levels on post-vaccination antibodies against influenza (45). Herein, subjects with vitamin D levels in the range of 25.68-32.99 ng/ml had a positive association with higher antibody titers when compared with subjects in the lower range of vitamin D in our population (4.1-18.89 ng/ml). The other 2 quartiles of vitamin D 18.9-25.67 and 33-69.8 did not show a statistically significant association with higher antibody titers but only a trend as seen in **Fig. 9**. In our study no information was available on whether our subjects were taking vitamin D supplementation and for how long. We just recorded vitamin D levels at a particular timepoint. Thus, it was not possible to know how long these levels were sustained and if there were previous fluctuations. The role of vitamin D in the antibody response after vaccination has not been systematically examined but future studies are warranted. It is worth mentioning that vitamin D supplementation has been tested even in hospitalized COVID-19 patients without definitive answers (46).

In conclusion, in the present study of 712 individuals we show that the antibody titer generated after 3 months from the last dose of COVID-19 vaccination is negatively associated with age, smoking and male gender after the age of 40 years. No association was found between antibody titer and obesity, diabetes, hypertension or use of statins. DHEAS levels were negatively correlated with age but did not appear to have any effect on the antibody titer. Vitamin D levels showed a positive association with antibodies titer. Moreover, by measuring the antibody titer in 198 of our subjects 3 weeks post-vaccination we concluded that there is a decline in the titer between 3 weeks and 3 months. Limitations of our study include the reliance on questionnaire for anthropometric measurements and the reporting of diabetes and hypertension. The final multiple linear regression model that was generated has an adjusted r^2^=0.14 (**Table 3**), meaning that this model explains roughly only 14% of the variance in our data and indicates that other parameters, besides the ones already taken into consideration, should be included so as to further improve the model. Advantages of our study include the relatively high number of participants, the strict criterion of excluding all subjects that at least one parameter (of those examined) is missing, the use of a multiple linear regression model taking into account all parameters examined in parallel. Last but not least, to the best of our knowledge, this was the first study to examine DHEAS and 25(OH)D levels as potential modulators of the SARS-COV-2 antibody titer response post-vaccination.

## Data Availability

Some or all data sets generated during and/or analyzed during the current study are not publicly available but are available from the corresponding author upon reasonable request.

## Acknowledgments

This study was funded by internal funds of Division of Endocrinology, University of Patras. We express our gratitude to Roche Diagnostics (Hellas) for donating the kits for quantitative determination of antibodies (including IgG) to SARS CoV 2 spike (S) protein receptor binding domain (RBD).

